# Complement and coagulation cascade cross-talk in endometriosis and the potential of JAK inhibitors – a network meta-analysis

**DOI:** 10.1101/2025.03.25.25324597

**Authors:** Monika Golinska, Aleksander Rycerz, Matylda Sobczak, Jedrzej Chrzanowski, Konrad Stawiski, Wojciech Fendler

## Abstract

**Background:** Lack of knowledge on the processes driving endometriosis hinders early detection and therapy development. Our purpose was to identify key molecular events involved in lesion formation across diverse populations and to detect transcriptomic changes in eutopic endometrium that accompany endometriosis.

**Methods:** We searched Gene Expression Omnibus and ArrayExpress and performed differential gene expression analysis and a network meta-analysis on nine qualifying datasets. Those contained transcriptomic data on: 114 ectopic endometrium samples (EL), 138 eutopic endometrium samples from women with endometriosis (EEM) and 79 eutopic endometrium samples from women without endometriosis (EH). Gene ontology and enrichment analysis was performed in DAVID, Metascape and Cytoscape and drug repurposing was done in CMap.

**Results:** EEM compared to EH upregulated CCL21 and downregulated BIRC3, CEL and LEFTY1 genes (|log2FC|>0.5, p<0.05). EL showed increased expression of complement and serpin genes (EL vs EEM: C7, logFC = 3.38, p <0.0001; C3, logFC = 2.40, p<0.0001; SERPINE1, logFC = 1.02; p<0.05; SERPINE2, logFC = 1.54, p<0.001) and mast cells markers (EL vs EEM: CPA3, logFC = 1.54, p<0.0001, KIT, logFC=0.74, p<0.001). Functional enrichment analysis highlighted complement and coagulation, inflammation, angiogenesis and ECM as drivers of endometriosis. Pharmacogenomic analysis indicated JAK, CDK and topoisomerase inhibitors as therapy targets.

**Conclusion:** Our results suggest an interplay between complement and coagulation, mast cells, ECM and JAK/STAT3 pathway in endometriosis. We underscore the significance of complement C3 and propose JAK inhibitors as therapy candidates. Detected expression differences between EEM and EH are important for the development of diagnosis via endometrial biopsy.

**WHAT IS ALREADY KNOWN ON THIS TOPIC:** Pathways and genes involved in endometriosis lesions formation are not well characterised. Studies encompassing diverse patients populations are missing.

**WHAT THIS STUDY ADDS:** This study reveals the transcriptomic profile of endometriosis, obtained via integration of nine different datasets spanning various ethnicities and demographics. It demonstrates the importance of complement and coagulation cascades, mast cells and JAK/STAT3 pathway in lesion development. Our meta-analysis identifies transcriptomic differences in eutopic endometrium of women with and without endometriosis which include changes in *CCL21*, *BIRC3*, *CEL* and *LEFTY1* expression.

**HOW THIS STUDY MIGHT AFFECT RESEARCH, PRACTICE OR POLICY:** Our comprehensive analysis of endometriosis transcriptomic profile highlights genes and pathways that should be explored further as disease biomarkers. JAK inhibitors currently used in clinic in other autoimmune diseases show treatment potential. Gene expression differences between eutopic endometrium of women with and without endometriosis should be further explored as biomarkers in endometrial biopsy.

## INTRODUCTION

Lack of knowledge on the key processes that drive endometriosis hinders its early detection and therapy development. There is a need to define those molecular events and to understand how they interact to foster lesion implantation and maintenance.

Endometriosis is a chronic and complex disease currently showing a median diagnostic delay of 7- 9 years^1,2^. There has been a significant progress in the development of endometriosis imaging protocols^3^, however, laparoscopy remains a gold standard for final diagnosis. There is a need to explore the less invasive endometrial biopsy option. To consider this strategy, the in-depth knowledge on the molecular differences in eutopic endometrium of healthy controls and women with endometriosis is needed.

Several attempts have been made at delineating disease biomarkers, but to date this has not yet proven successful. Various omics technologies enabled identification of key genes related to the pathophysiology of endometriosis. However, a consensus has not yet been reached, and we are still missing the focal points on which to concentrate the therapeutic endeavors. A multi-cohort analysis is needed to address the issue in an unbiased and comprehensive manner.

In this article we aimed to better understand complex events that underlie endometriotic lesion formation and progression. To achieve this, we systematically reviewed endometriosis data and performed network meta-analysis on chosen datasets. We generated a transcriptomic profile of endometriosis, determined the key pathways involved in lesion formation and explored possible drug candidates for endometriosis therapy.

## METHODS

### Search strategy and study selection

Gene Expression Omnibus (GEO) and ArrayExpress were searched using terms “endometriosis” and “Homo sapiens” and filtered with terms “Expression profiling by array” or “Expression profiling by high throughput sequencing”. MEDLINE/Pubmed was searched for publications that correspond to those publicly deposited datasets.

Studies included in the analysis had to contain at least two tissues of interest: ectopic endometrium - endometrial lesion (EL), eutopic endometrium from women without endometriosis (EH) or eutopic endometrium from women with endometriosis (EEM).

Inclusion criteria were predefined and stringent to minimize the risk of bias, focusing on datasets using RNA-Seq or microarray technologies, with available raw data. Transcriptomic analysis had to be performed directly on human endometrial tissue, that had not been subjected to any manipulation or cell isolation prior to RNA extraction. Samples had to be taken from patients not on hormonal treatment in three months preceding tissue collection. The presence or absence of endometriosis had to be confirmed with laparoscopy for samples to be included in our study. Only datasets with accompanying publication were considered to ensure all information about samples was available. Datasets with incomplete information were excluded to reduce variability and minimize errors. A full list of inclusion / exclusion criteria together with the PRISMA selection flowchart are summarized in Table S1 and Fig. 1. Two independent reviewers screened datasets for relevance, and any discrepancies were resolved in discussion with a third reviewer. PRISMA guidelines were followed, and study protocol was registered in PROSPERO (ID CRD42024548098).

**Figure 1.**
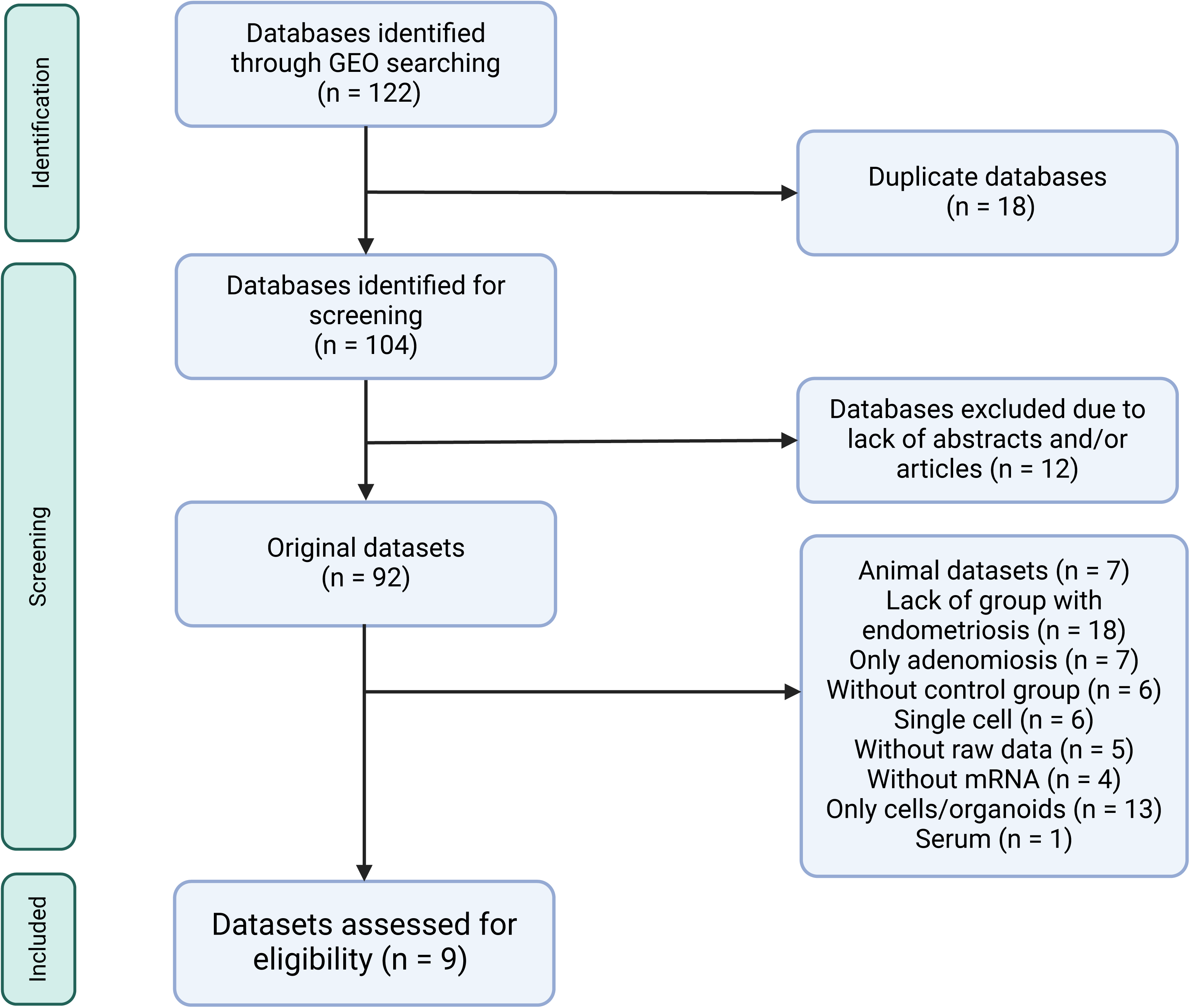
PRISMA flow diagram of Gene Expression Omnibus search for transcriptomic data comparing eutopic and ectopic endometrial tissue. All datasets from ArrayExpress were also deposited in Gene Expression Omnibus thus they were not further considered in the selection process.

### Data extraction and Differential Gene Expression

Each dataset was analyzed individually to ensure that data-specific preprocessing and normalization steps were applied appropriately. For microarray datasets, the raw data files were retrieved from the GEO repository using the R package “GEOquery”. The preprocessing of microarray data was conducted following the manufacturer’s protocols. Background correction and quantile normalization were applied for all array data. To adjust for differences in library size and transcript length, the raw reads count from RNA-Seq datasets were normalized and scaled using the average transcript length for each sample. Following this, library size normalization was performed using the Trimmed Mean of M-values (TMM). After preprocessing, group comparisons were conducted on the normalized datasets to identify differentially expressed genes (DEGs) between experimental groups. For this purpose, the “limma” package in R was utilized. The analysis generated log fold change (logFC) values and their corresponding standard error (SE) values which were used for further analysis.

### Network meta-analysis

Network meta-analysis on gene expression was performed using “netmeta” package. Although meta-analysis allows for the determination of both direct and indirect effects, in our subsequent analyses, we focused on the combined effect to maximize the quality of the analyzed data and reduce the influence of less reliable direct or indirect effects. For investigated difference measurement we used logFC and its corresponding standard error. These were interpreted as the mean difference and the standard error of the mean difference, respectively, which are widely used metrics in comparative gene expression studies. This standardization ensures that the results are both interpretable and comparable across datasets.

We performed 7664 network meta-analysis for genes that occurred in each of datasets included in the study. Genes with a p-value < 0.05 were considered statistically significant and a |logFC|>0.5 was used to filter genes with biologically meaningful changes in expression.

### Risk of bias

To reduce the risk of bias, we included studies with raw data deposited and results published in peer-review journals. Information from accompanying publication was used to ascertain the quality of the study and to identify if the absence of endometriosis was properly determined and to confirm that tissue did not undergo any manipulation prior to RNA isolation.

Heterogeneity was evaluated using I² statistics and Cochran’s Q-test, while sensitivity analyses validated the robustness of findings. Funnel plots were generated to assess bias. These measures ensured a thorough evaluation of potential biases, enhancing the reliability and validity of the meta-analytic findings.

### Gene ontology and pathway analysis

The list of DEGs obtained from the network meta-analysis was submitted to DAVID for gene ontology and KEGG and Reactome pathways analysis. For functional clustering, we applied a cut-off enrichment score of >2.5, p<0.05 and medium classification stringency. The same list of DEGs was analyzed in Metascape v3.5.2024.0101 and the most enriched terms were visualized in Cytoscape v3.10.2.

### Computational pharmacogenomics

To identify pharmacological compounds likely to reverse endometriosis gene signature, we queried drug repurposing reference database - CMap. We submitted a list of 150 up- and 150 down-regulated genes that had the highest combined logFC for EL vs EH and EL vs EEM comparisons and p<0.05.

## RESULTS

### Characteristics of chosen studies

Nine datasets met the inclusion criteria (Fig. 1 and Table S1) and were included in the analysis (Table 1). Those contained transcriptomic data on 114 ectopic endometrium samples (EL), 138 eutopic endometrium samples from women with endometriosis (EEM) and 79 eutopic endometrium samples from women without endometriosis (EH). The absence of endometriosis in the healthy (EH) group had to be confirmed during laparoscopic procedure. Tissues were collected at three different continents and encompassed all types and stages of endometriosis (clinical data in Table S2).

**Table 1.**
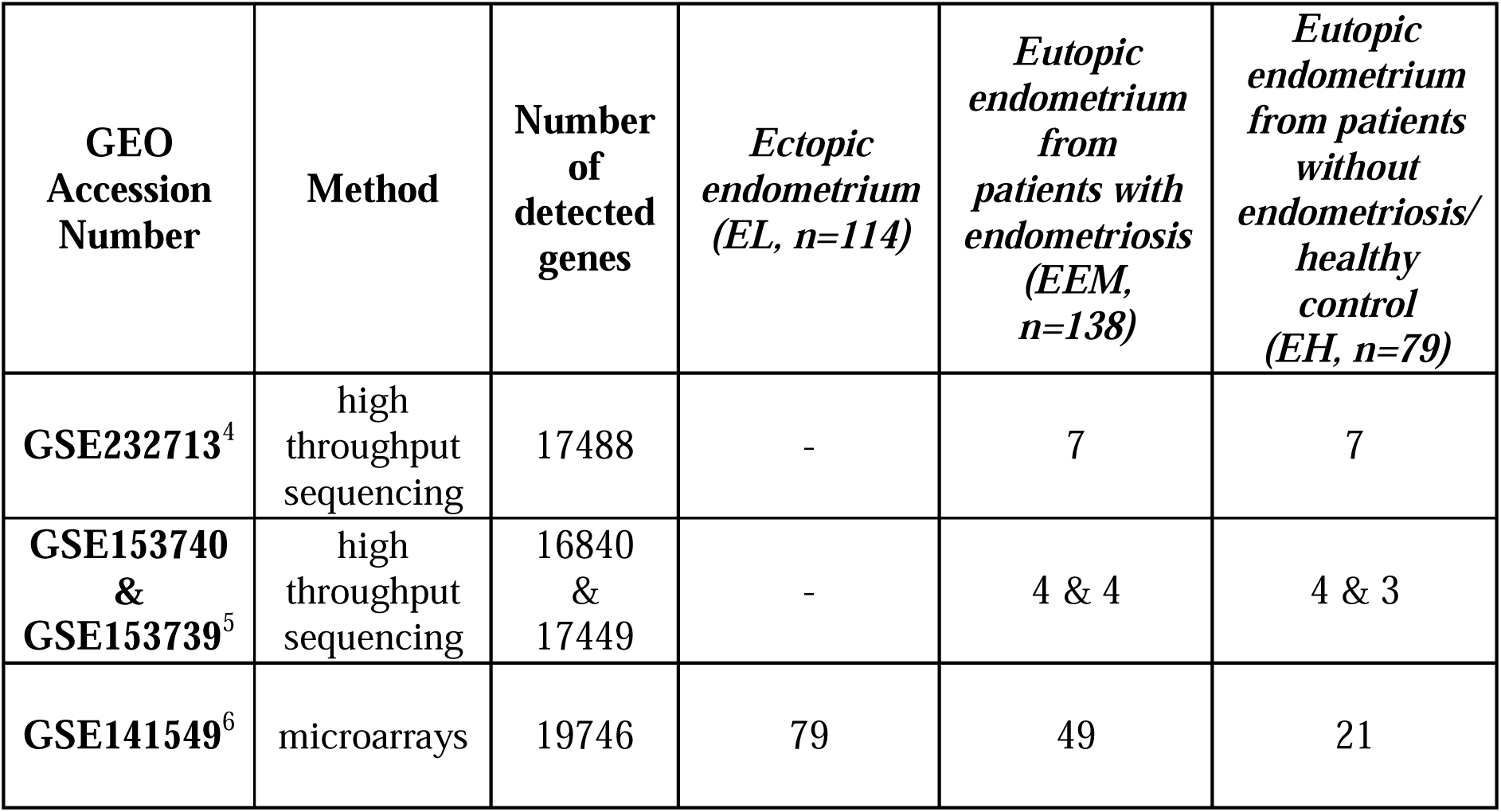
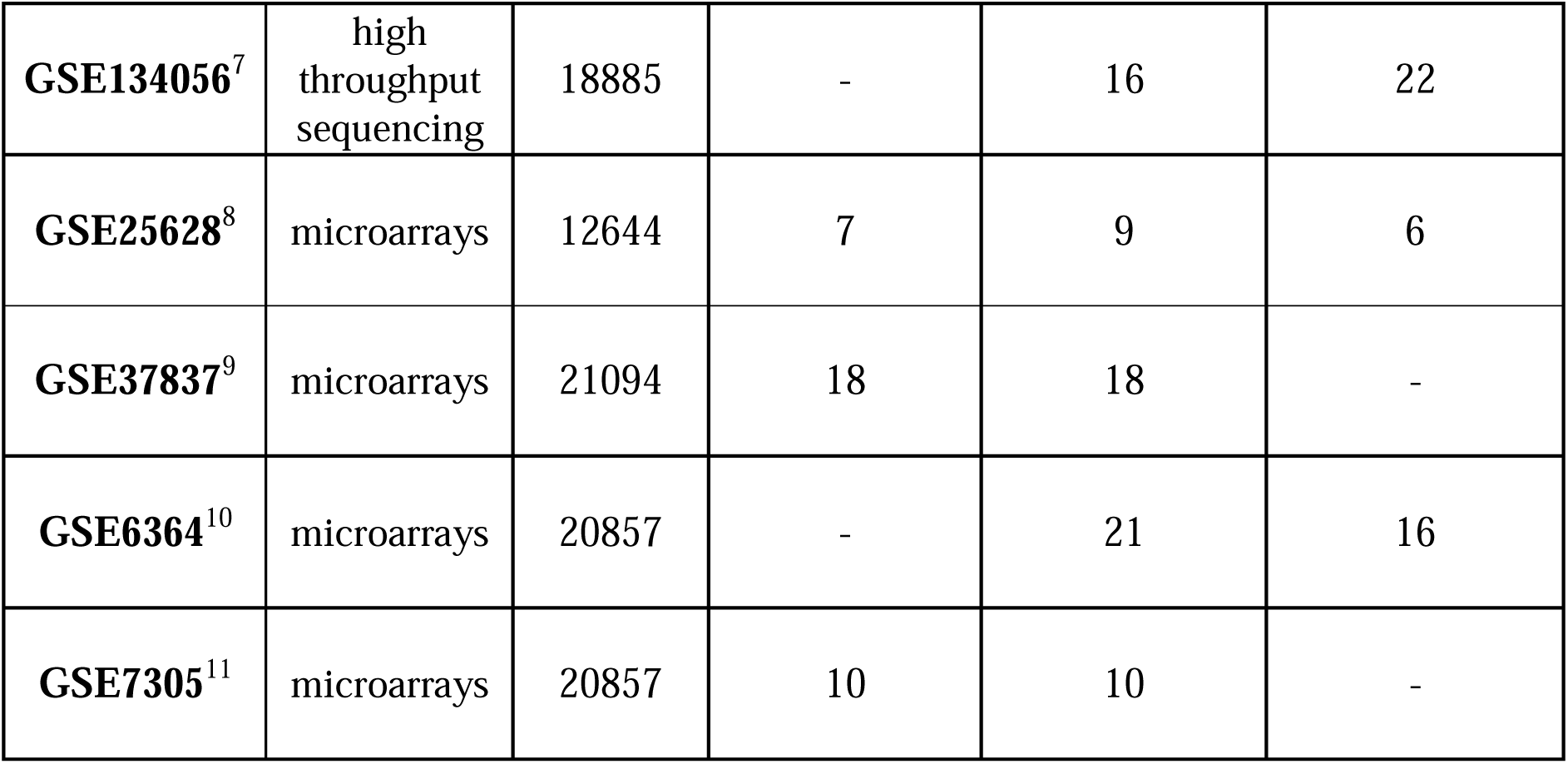
Characteristics of GEO datasets chosen for the network meta-analysis.

| **GEO Accession Number** | **Method** | **Number of detected genes** | ***Ectopic endometrium (EL, n=114)*** | ***Eutopic endometrium from patients with endometriosis (EEM, n=138)*** | ***Eutopic endometrium from patients without endometriosis/ healthy control (EH, n=79)*** |
| --- | --- | --- | --- | --- | --- |
| **GSE232713**<sup>4</sup> | high throughput sequencing | 17488 | - | 7 | 7 |
| **GSE153740 & GSE153739**<sup>5</sup> | high throughput sequencing | 16840 & 17449 | - | 4 & 4 | 4 & 3 |
| **GSE141549**<sup>6</sup> | microarrays | 19746 | 79 | 49 | 21 |
| **GSE134056**<sup>7</sup> | high throughput sequencing | 18885 | - | 16 | 22 |
| **GSE25628**<sup>8</sup> | microarrays | 12644 | 7 | 9 | 6 |
| **GSE37837**<sup>9</sup> | microarrays | 21094 | 18 | 18 | - |
| **GSE6364**<sup>10</sup> | microarrays | 20857 | - | 21 | 16 |
| **GSE7305**<sup>11</sup> | microarrays | 20857 | 10 | 10 | - |

References next to each dataset identifier refer to the original publication.

### Transcriptomic profile of eutopic and ectopic endometrium

Differential expression analysis was performed for each of the three comparisons: EL vs EEM, EL vs EH and EEM vs EH. Using p <0.05 and |logFC|>0.5, we identified 1109 DEGs between EL and EEM, 1267 DEGs between EL and EH and 4 DEGs between EEM and EH (Fig. 2). The heatmap of top 40 up and down regulated genes for all comparisons per dataset is presented in Fig. 2D. The full list of network meta-analysis results is deposited in Supplementary Dataset S1.

**Figure 2.**
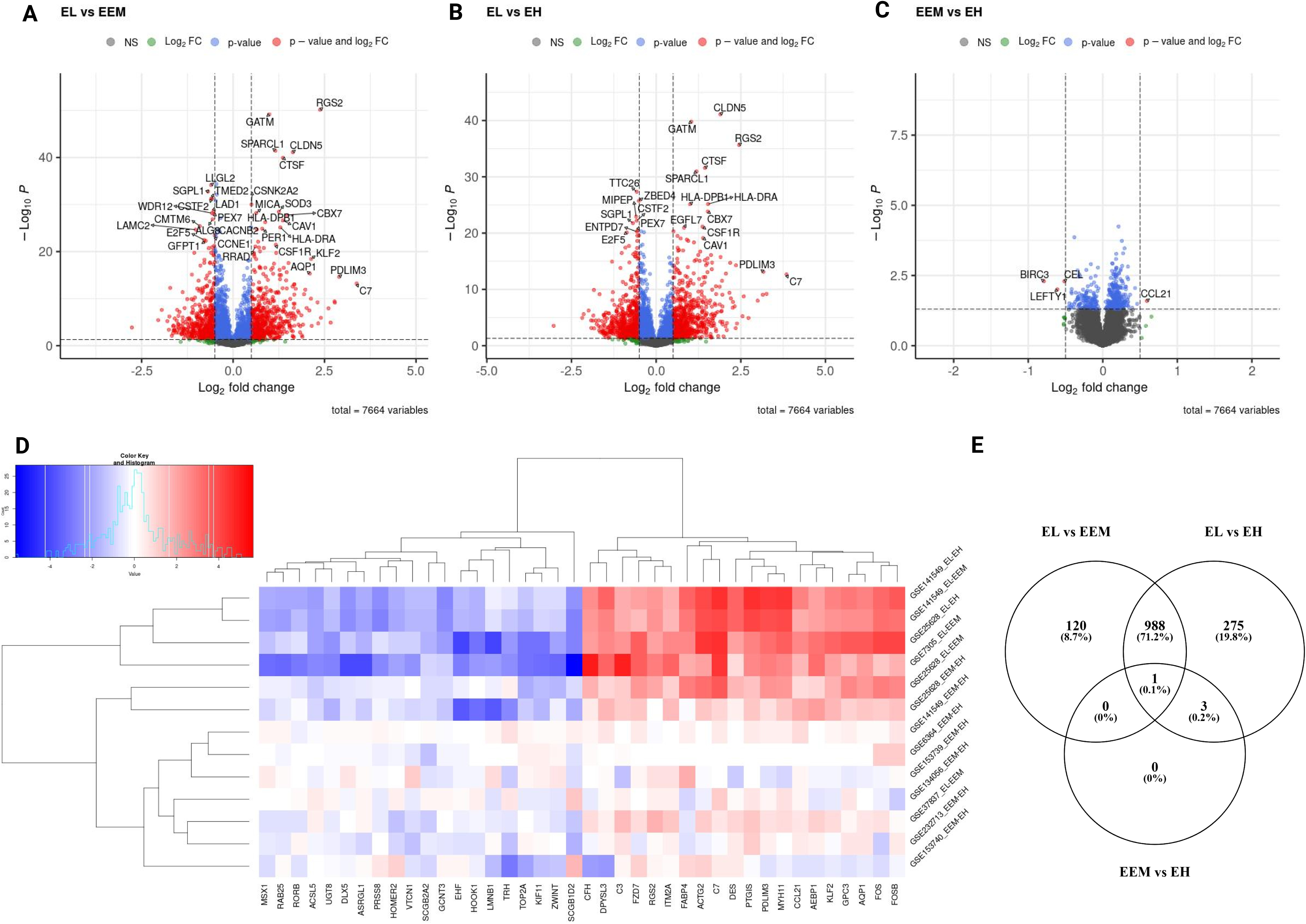
Differentially expressed genes identified by network meta-analysis. Volcano plots showing differentially expressed genes for the following comparisons (A) endometriotic lesions versus endometrium from women with endometriosis, (B) endometriotic lesions versus endometrium from women without endometriosis, (C) endometrium from women with and without endometriosis. For graphs A to C, red points signify genes with logFC less than - 0.5 or more than 0.5 and p-value less than 0.05, blue points signifiy genes with logFC belonging to -0.5 to 0.5 range and p-value less than 0.05, green points signify genes with logFC less than -0.5 or more than 0.5 and p-value more than 0.05 and grey dots signify genes with with logFC belonging to -0.5 to 0.5 range and p-value more than 0.05. Genes with p- values<1×10^−6^ and log2FC>|2| (Fig. 2A-B) and p-values<0.05 and log2FC>|0.5|(Fig. 2C) are labelled. Heatmap with top 40 most differentially expressed genes per comparison per dataset (D). Expression pattern of differentially expressed genes per comparison type (E). EL – endometrial lesion, EEM - eutopic endometrium from women with endometriosis, EH – eutopic endometrium from women without endometriosis.

Meta-analysis revealed that transcriptomic profile of lesions was profoundly different from that of eutopic endometrium (Fig. 2A-B) while the eutopic endometrium from women with (EEM) and without endometriosis (EH) differed in the expression of four genes only (Fig. 2C). *BIRC3*, *CEL* and *LEFTY1* were significantly less expressed in endometrium of women with endometriosis than without (logFC = -0.79, p = 0.0051; logFC = -0.52, p = 0.0051; logFC = -0.61, p = 0.0099, respectively). *CCL21* was significantly higher in EEM versus EH (logFC=0.59, p = 0.0255) and even higher when EL with EEM was contrasted (logFC=1.57, p < 0.0001, Fig. 2C). C-C motif chemokine ligand 21 (*CCL21*) is an inflammatory mediator associated with moderate to severe endometriosis^12^, however to-date its use as a disease biomarker has failed. Our results showed a directional increase of *CCL21* from endometrium of healthy patients through that of endometriosis sufferers to lesions themselves indicating its role in the eutopic endometrium inflammation in patients with endometriosis.

### Pathways contributing to lesion development

In further analysis, we selected genes that showed differential expression in both EL vs. EEM and EL vs. EH comparisons (the intersection of the sets, Fig. 2E) and exhibited the same direction of expression. For p<0.05 and |logFC|>0.5 we obtained a list of 989 DEGs: 536 upregulated and 453 downregulated, on which we performed functional annotation and enrichment analyses (Fig. 3A and detailed in Table S3A). Results presented below satisfied a p value below 0.0001. Those analyses revealed that most biological processes involved in the formation of endometriotic lesions were linked to cell adhesion (6.6%), inflammatory response (5.5%) and regulation of angiogenesis (2.9%). The gene ontology molecular functions analysis showed that the DEGs were significantly enriched in protein binding (76.7%), identical protein binding (15%) and extracellular matrix structural constituent (2.5%). In the cellular component, DEGs were mainly involved in extracellular exosome (21.2%), extracellular region (21.2%) and extracellular space (17.4%).

**Figure 3.**
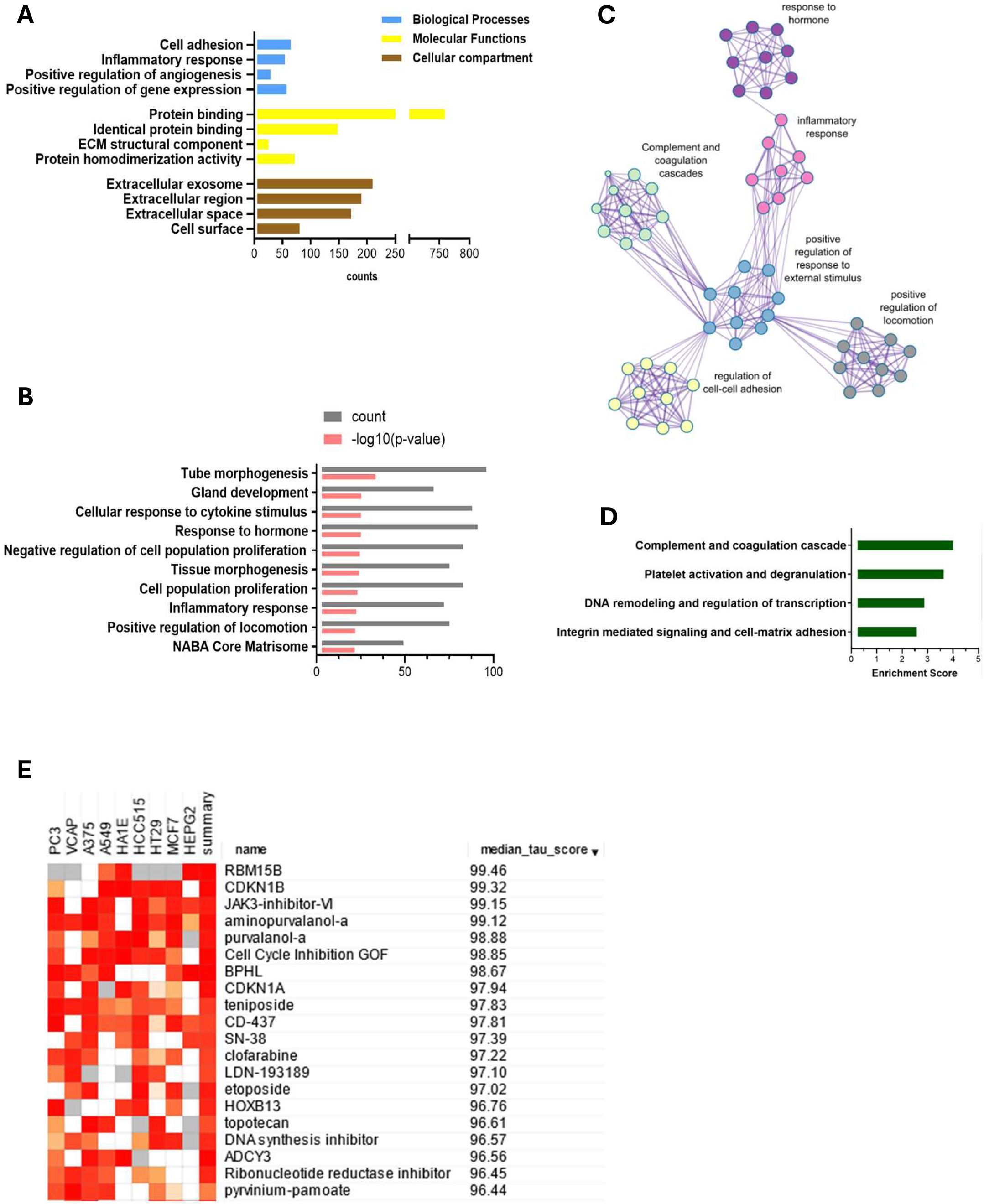
Enriched pathways analysis, functional clustering and computational pharmacogenomics of DEGs between endometriosis lesions and eutopic endometrium. Gene ontology analysis using DAVID (A) reveals the importance of inflammation, cell adhesion, angiogenesis and ECM remodeling. Metascape enrichment analysis (B) and relationship network of enriched terms visualised in Cytoscape (C) show key events that contribute to endometriosis development. Those include inflammatory and hormonal response and proliferation and locomotion. Functional annotation clustering reports the highest enrichment score for complement and coagulation cascade, platelet activation, DNA remodeling and integrin mediated signaling respectively (D). Top 15 drug candidates identified using a drug repurposing reference database – CMap. and showing median tau value above 95. JAK, CDK and topoisomerase inhibitors are identified as potential pharmacological targets for endometriosis therapy (E).

KEGG analysis showed enrichment in complement and coagulation cascades (2.5%), Staphylococcus aureus infection (2.3%) and cell adhesion molecules (2.7%). The analysis against Reactome database revealed a key role of extracellular matrix organization (5.2%), regulation of complement cascade (1.5%) and complement cascade (1.6%) (Table S3B).

A further pathway enrichment analysis was performed with Metascape (Fig. 3B and Table S4) and visualized in Cytoscape (Fig. 3C). Tube morphogenesis, which relates to vascular development, was the highest ranked result of enrichment analysis (Fig. 3B). Inflammatory and hormonal response as well as locomotion and proliferation were amongst the top 10 most enriched pathways with count value above 75. Functional annotation clustering revealed that the complement cascade was the most enriched with the score of 4.14, followed by platelet activation pathways, DNA remodeling and regulation of transcription and cell/cell-matrix adhesion processes (enrichment score of 3.77, 3.02 and 2.71 respectively (Fig. 3D).

### Altered expression of complement and coagulation pathway genes

Complement and coagulation cascade was the most enriched KEGG pathway for the ectopic versus eutopic endometrium comparison (Fig. 3D). Genes including *C3*, *C2*, *C3* and *SERPIN* superfamily genes involved in this pathway were amongst the most differentially expressed in endometrial tissue (Fig. 4 and Dataset S1).

**Figure 4.**
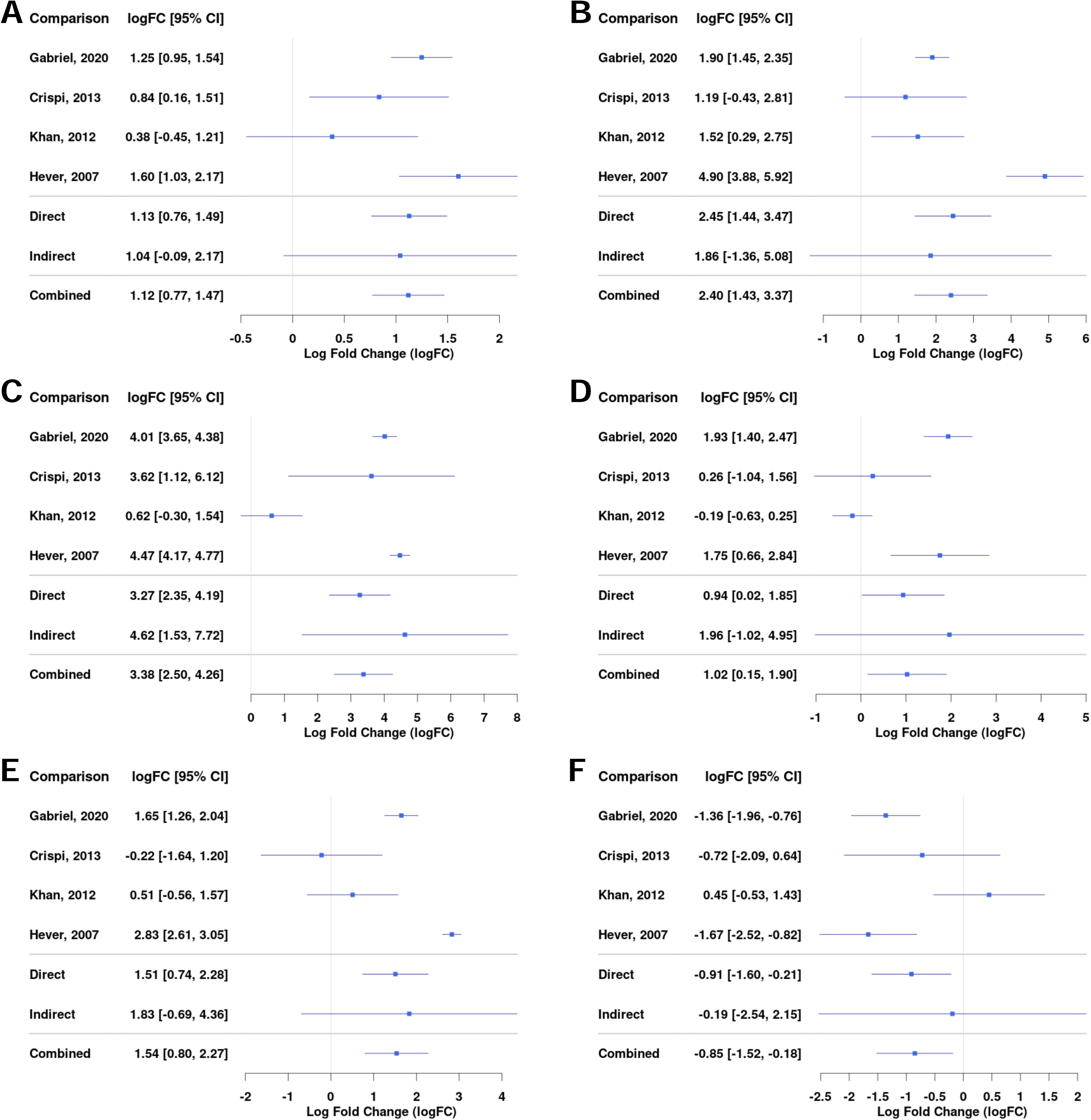
Differential gene expression across studies for selected genes from the complement and coagulation pathway for the comparison between EL vs EEM. Forest plot showing the expression of *C1QA (complement C1q A chain)* - A, *C3 (complement C3)* - B, *C7 (complement C7)* - C, *SERPINE1 (serpin family E member 1)* - D, *SERPINE2 (serpin family E member 2)* - E, *SERPINA5 (serpin family A member 5)* - F. Direct and indirect comparisons from meta-analysis are presented in row number five and six. The indirect comparisons had a low impact on the combined comparison outcome due to the analyses being performed on datasets containing comparisons between EL and EEM (Table 2).

Complement genes *C1QA* (logFC = 1.12; 95%CI = 0.77, 1.47), *C3* (logFC = 2.40; 95%CI = 1.43, 3.37) and *C7* (logFC = 3.36; 95%CI = 2.50, 4.26) were upregulated in endometrial lesions and showed high logFC values (Fig. 4A-C). *C7* was the gene that showed the highest level of upregulation among all examined genes.

Serpins regulate coagulation fibrinolysis processes^13^ and were implicated in the development of endometriosis^14–16^. Our network meta-analysis showed that serpin genes were differentially expressed between endometrial lesions and eutopic endometrium. In comparison with the above presented complement genes, serpin family genes were characterized by more heterogenous expression between investigated datasets. *SERPINE1* and *SERPINE2* were upregulated (logFC = 1.02; 95%CI = 0.15, 1.90 and logFC = 1.54; 95%CI = 0.80, 2.27 respectively) while *SERPINA5* was downregulated in lesions (logFC = - 0.85; 95%CI = -1.52, -0.16, Fig. 4D-F). Detailed comparisons for each of the subgroups can be found in Dataset S1.

### Mast cells markers

Our data showed an upregulation in the expression of mast cells markers including *CPA3* (logFC = 1.54; 95%CI = 0.96, 2.11), *KIT* (logFC = 0.74; 95%CI = 0.30, 1.18), *MS4A6A* (logFC = 0.71; 95%CI = 0.30, 1.11) and markers of mast cells activation *FCGR2B* (logFC = 0.78; 95%CI = 0.20, 1.35) and *S100A10* (logFC = 0.87; 95%CI = 0.38, 1.35, Fig. S3A-E).

The expression of *MS4A4A* and *MS4A2* was also higher in lesions (Dataset S1). Higher amounts of mast cells and their increased degranulation have been reported in endometrial tissue of animal models and humans^17,18^; mast cells colocalized to the vasculature of ovarian endometriomas and they were found to promote endometrial cells migration in in vitro assays^19^.

### Repurposing JAK and CDK inhibitors for endometriosis therapy

We used CMap drug repurposing software to find most probable connections between therapeutic drugs and our network meta-analysis results. A median tau score value of 90 or above is considered the typical threshold for assessing meaningful drug-induced effects. We applied a median tau score cutoff at 95 and selected the top 15 hits. This analysis indicated that the candidates most likely to reverse the endometriosis mRNA profile were cyclin-dependent kinase (CDK) inhibitors, JAK and topoisomerase inhibitors (Fig. 3E).

JAK/STAT3 pathway is thought to govern migratory and invasive properties of cells. Its prolonged activation in breast cancer was linked with tumor development and resistance to taxane and platinum therapy^20^. Our results showed an increase in the expression of *STAT5A* (logFC = 0.83; 95%CI = 0.57, 1.08) and *STAT5B* (logFC = 0.57; 95%CI = 0.37, 0.77) in lesions compared with control tissue (Fig. S3 G-H). *JAK3* significantly increased as well but logFC value was below 0.5 (Dataset S1).

JAK inhibitor Ruxolitinib reduced epithelial ovarian cancer cell viability and caused growth inhibition of Tam resistant breast cancer cells *in vitro*. It was shown to lower mRNA VEGF expression and reduce the number of vessels and overall tumor weight in chorioallantoic assay^20^. Ruxotinilib is currently tested in combination therapy for endometrial cancer but its use *in vitro* or in preclinical models of endometriosis has not been reported. Tofacitinib, another JAK inhibitor, showed a decrease in endometrial lesion size in mice and reduced proliferation of endometrial cancer cells *in vitro*^21^.

## DISCUSSION

Understanding the main pathways involved in endometriosis development is necessary for the successful biomarker discovery and improved therapy outcomes. Combining data in meta-analysis, we highlight pathogenetic mechanisms that are critical for lesion formation regardless of endometriosis subtypes and patients’ characteristics.

### Endometrium of women with endometriosis differs from healthy controls

We detected differences in gene expression between endometrium of healthy women and those suffering from endometriosis thus showing that endometriosis can also affect eutopic endometrium (Fig. 2C). *CCL21* was upregulated whilst *BIRC3*, *LEFTY1* and *CEL* were downregulated in EEM versus EH. Increased expression of *CCL21* could suggest that this gene takes part in inducing early inflammatory changes in eutopic endometrium in women with endometriosis and that it continues its role in established lesions (Dataset S1). Baculoviral IAP repeat containing 3 (*BIRC3*) has not been studied in the context of endometriosis. However, its mutations are often present in endometroid adenocarcinoma and endometrial cancer^22^. In the latter, the lower protein levels of *Birc3* correlate with worse patient survival. One could speculate that the decreased *B*irc3 expression in EEM could contribute to the transformation of endometrium into lesions. Endometrial bleeding associated factor (*EBAF*/*LEFTY1*) partakes in the regulation of cyclical exfoliation of endometrium and in decidualization. Healthy endometrium does not express *LEFTY1* during implantation window while endometrium of women suffering from endometriosis as well as infertility showed its expression^23^. Our results agree with that finding and suggest that the higher *LEFTY1* expression in EEM group could contribute to endometriosis-related infertility. *CEL* gene encodes carboxyl ester lipase, which partakes in cholesterol and lipid-soluble vitamin ester hydrolysis. Its role so far is implicated in diabetes and hereditary pancreatitis and progression of atherosclerosis. The *CEL* gene has not yet been studied in the context of endometriosis.

### The complement and coagulation cascade in lesion formation

We further focused on delineating the expression profile that can differentiate ectopic endometrium from eutopic endometrium from women with and without endometriosis (Fig. 2). Gene ontology analyses highlighted crucial events accompanying lesion formation. Those were immune system activation, angiogenesis, regulation of transcription, response to hormones and cytokines, cell adhesion and ECM – cell surface interactions (Fig. 3). The importance of immune system deregulation in endometriosis has been reported previously; various inflammatory phenotypes have been associated with increased risk of endometriosis^24,25^. Our result showed that the complement system and platelet coagulation are the two most enriched pathways in endometriosis (Fig. 3D). Both processes are essential in natural endometrium growth and shedding cycle. The fact that both pathways are the most enriched agrees with the current theory that women prone to endometriosis are likely to have a different, dysregulated peritoneal microenvironment. The complement system is a mediator of tissue growth and regeneration and its activation has for a long time been implicated in the development of autoimmune disease and in promoting tumor growth. Its dysregulation could therefore provide means for immunosurveillance escape and facilitate the implantation of lesions. Its importance in the development of endometriosis has been suspected since the 80’s^26^ and confirmed more recently^27,28^. Higher amounts of C1, C3 and C5 have been detected in serum^29^ and peritoneal fluid of women with endometriosis^30,31^. Various complement proteins were shown to be present in epithelial cells of endometrial lesions and ovarian cancer tumors. Its local synthesis and deposition has been correlated with progression of various cancer types.

Our data revealed an increased mRNA expression of *C1q*, *C2*, *C6* but especially *C3* and *C7* in the lesions (Fig. 4A-C, Dataset S1). *C7*, a complement cascade member responsible for initiation of membrane attack complex, was the most overexpressed gene with the highest fold change in our comparison between diseased and control tissue suggesting its significant role in lesion formation (Fig. 2A-B, Fig. 4C). C7 was found to contribute to inflammation and tissue damage in endometriosis^32^; it has previously been shown overexpressed in ovarian cancer^25^ and stromal cells of endometriomas^33^.

C3, a major effector, at which all complement pathways converge, was one of the most differentially expressed genes in endometriosis (Fig. 4B, Figure S2). C3 dysregulation is involved in most if not all inflammatory diseases; it has been found upregulated in cancer, cardiac and neurological diseases, asthma and obesity. Patients with inflammatory bowel disease had a higher expression of C3 in their intestinal tissue and this was thought to contribute to chronic inflammation and tissue injury. A similar situation could occur in endometriosis; increased C3 expression could contribute to inflammation-driven peritoneal tissue injury, which in turn would facilitate lesion implantation. Glandular epithelial cells found in endometrial lesions were shown to produce C3 locally^34^. The activity of both complement cascade members C3 and C4 was higher in serum of women with endometriosis than those without^35^. Increased amounts of C1, C3 and C5 were detected in serum^29^ and peritoneal fluid of women with endometriosis^30,31^. Similarly, in lesion-bearing mice, C3 was increased in their peritoneal fluid. Animals with *C3* knockdown formed smaller endometrial cysts and on average less of them^34^.

C3 seems pivotal to endometriosis pathology and given its strong upregulation and presence both in tissue (Fig. 4B) as well as peritoneal fluid of endometriosis-sufferers, it poses an interesting target for early diagnosis and therapy. It has already been proposed as an endometriosis serum biomarker. Since in gastric cancer C3 tissue deposition correlated negatively with plasma levels^36^, further research is needed to confirm C3 suitability as endometriosis biomarker. As far as treatment is concerned, C3 inhibitors have entered clinical trials in anti-ovarian cancer therapy^37^ and treatment against inflammatory bowel disease. Our results indicate that biomarker and therapeutic potential of C3 should be studied in endometriosis in more depth.

Our results revealed strong enrichment in coagulation cascade and showed a dysregulation of *SERPIN* superfamily genes in endometrial lesions (Fig. 3D, Fig. 4D-F, Fig. S4) suggesting an imbalance in the coagulation-fibrinolysis processes^13^.

*SERPINE1* and *SERPINE2* were increased in endometrial lesions (Fig. 4D-E). *SERPINE1-* encoded PAI-1 was found increased in deep infiltrating lesions^38^ and correlated with ovarian cancer proliferation and overall poor prognosis^39^. PAI-1 inhibition resulted in decreased lesion size^40^. *SERPINE2* was implicated in modulating DNA damage response and favoring cancer cell invasion^41^. Its pro-metastatic activity has been linked to extracellular matrix remodeling and increase in matrix metalloproteinase 9 (MMP-9) expression^42,43^.

Reduction in *SERPINA5* expression was linked with an aggressive tumour phenotype and poor prognosis in endometrial and ovarian serous carcinomas, in the latter it was correlated with downstream activation of MMP9^44^. Our meta-analysis revealed lower *SERPINA5* and higher *MMP9* expression in endometrial lesions (Fig. 4F, Fig. S3F). Moreover, ECM interactions were indicated in enrichment analysis (Fig. 3A-B, D). Taken together, our results suggest that the imbalance in coagulation pathway may be affecting extracellular matrix remodeling and contributing to metastatic-like potential of endometriotic cells; thereby promoting lesion formation.

### JAK/STAT3 pathway inhibition

Our search for associations between endometriosis gene signature and CMap reference perturbagens highlighted the role of inhibitors of JAK, CDK and topoisomerase as possible therapy candidates (Fig. 3E). Our meta-analysis revealed an increased expression of both *STAT5A* and *STAT5B* in lesions compared with control tissue (Fig. S3 G-H).

Interestingly, increased C3 expression was shown to trigger JAK2/STAT3 pathway in gastric cancer, which led to subsequent increase of cell proliferation. C3 inhibition with CR1 decreased that activation^36^. Our results present a similar picture, complement C3 as well as JAK/STAT3 pathway seems to play a role in the development of endometriosis. This association needs further investigation. JAK inhibitors are already used in clinic for other autoimmune disease therefore their repurposing should be further tested for endometriosis therapy application.

### Proposed pathways crosstalk in endometriosis

It has been proposed that both complement system and coagulation pathways are tightly linked; coagulation factors have been reported to cleave and activate complement members C3 and C5^45^. On the other hand, C3 was shown to protect clots from fibrinolysis^46^. Increased amounts of C3 protein were shown to provoke mast cells activation and various mast cell mediators were implicated in the regulation of coagulation and fibrinolysis in anaphylaxis^47^. Our meta-analysis revealed that endometrial lesions had a higher expression of mast cell markers including *KIT*, *CPA3* and *MS4A6A, FCGR2B* and *S100A10* (Fig. S3A-E). An increased mast cells burden was detected previously in animal and human endometrial tissue^19^. Moreover, our results showed that endometrial lesions had a higher level of *STAT5A* and *STAT5B* (Fig. S5G-H), members of JAK/STAT3 pathway, which regulate mast cells^48^. Targeting mast cells with JAK inhibitors for alleviation of symptoms of endometriosis has been proposed almost two decades ago^49^ but not much research has been carried out on the topic since. Our current results fill this gap and suggest the use of JAK inhibitors as immunomodulators in endometriosis. Interestingly, a cooperation between mast cells, complement and coagulation pathways has been reported in an inflammatory disease - chronic spontaneous urticaria^50^. Our analysis indicates that there exists an interplay between complement and coagulation pathway, mast cells activation, ECM remodeling and JAK/STAT3 pathway (summarized in Fig. 5). To the best of authors knowledge, this relationship has not yet been studied in endometriosis and our results warrant a further in-depth look into those processes.

**Figure 5.**
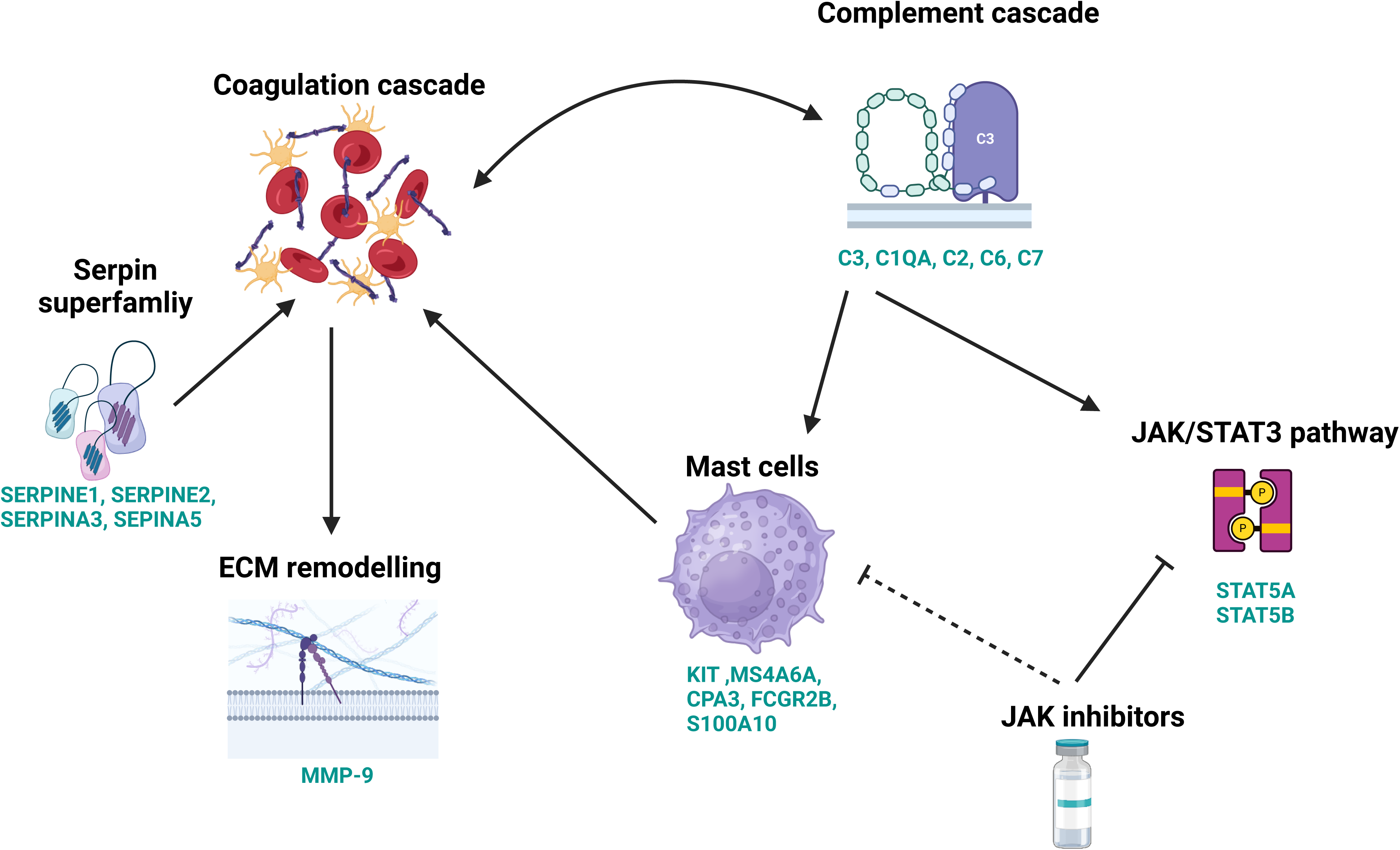
A schematic showing key molecular processes contributing to the development of lesions. An interplay between complement and coagulation pathway further influences mast cells activation, ECM remodelling and JAK/STAT3 pathway. JAK inhibitors carry potential for endometriosis therapy. Genes names in green signify differentially expressed genes for the comparison between EL and EEM. Created in BioRender.com.

## Strengths and Limitations

Our network meta-analysis enabled us to arrive at a consensus endometriosis signature. The use of publicly deposited endometriosis transcriptomic data collected at three different continents, spanning various ethnicities, age groups as well as various types and stages of endometriosis enabled a comprehensive, unbiased and multi-demographic comparison of endometriotic and control tissue.

The following limitations should be considered when interpreting our results. Our meta-analysis included only nine datasets because most of the studies lacked a control group, included therapeutic intervention or performed RNA isolation on processed tissue. Secondly, only published studies, where the absence of endometriosis was excluded by laparoscopy, were included in this meta-analysis. Therefore, publication bias may have occurred although none was indicated by the funnel plot.

## Conclusions and Clinical Implications

We highlight the role of complement and coagulation cascade in endometriosis and propose an interplay between both those processes and mast cells, ECM interaction and JAK/STAT3 pathway that need further investigation. We underscore the significance of C3 and call for further research into its diagnostic and therapeutic potential. Furthermore, we propose JAK inhibitors discovered in drug repurposing analysis and validated *in vitro*, as potential therapy candidates.

Our results show differences in expression in eutopic endometrium from patients with and without endometriosis. Those should be further explored to understand if they contribute to endometrial seeding. Detected gene differences may be potential biomarkers that could be used in the less invasive endometriosis biopsy and should be further studied.

## Data Availability

All data produced in the present study are available upon reasonable request to the authors

## Acknowledgements

This research is part of the project No. 2022/47/P/NZ5/02484 co-funded by the National Science Centre and the European Union Framework Programme for Research and Innovation Horizon 2020 under the Marie Skłodowska-Curie grant agreement No. 945339. For the purpose of Open Access, the author has applied a CC-BY public copyright licence to any Author Accepted Manuscript (AAM) version arising from this submission;”.

## Author contributions

M.A.G, W.M.F and conceived and supervised the study, A.R., M.S. performed GEO database search, M.A.G., A.R. and M.S. performed study selection, K.S. contributed to database search, A.R., J. C. performed differential genes expression, A.R., K.S. performed network meta-analysis, M.A.G performed GO and enrichment analysis, M.A.G performed pharmacogenomic analysis and cell culture in vitro experiments, M.A.G. and AR conducted quality control of the data, M.A.G. obtained study funding, M.A.G. and A.R drafted the manuscript, and W.M.F, M.A.G. and A.R revised the manuscript.

