## Supplement for "Complement and coagulation cascade cross-talk in endometriosis and the potential of JAK inhibitors – a network meta-analysis"

**Supplementary Tables and Figures:**

**Inclusion criteria**

Transcriptomic datasets containing endometriosis and a control group

Accompanied by the corresponding publication

The presence or absence of endometriosis confirmed via laparoscopic procedure

Patients not undergoing hormonal treatment three months prior to tissue collection

Transcriptomic analysis performed directly on endometrial human tissue

**Exclusion criteria**

Tissue processing post excision (for further cell isolation or single cell analysis)

Adenomyosis included as endometriosis group

ScRNA-seq analysis

Lack of whole genome mRNA sequencing and or profiling

Sequncing or profiling performed on cells or organoids isolated from tissue

Lack of raw data deposited in GEO

**Supplementary Table S1.** I**nclusion and exclusion criteria for selection of GEO datasets.**

| **GEO**  **Accession**  **Number** | **Author** | ***Age:***  ***mean***  ***(range)*** | ***Type of endometriosis*** | ***Stage of endometriosis*** | ***Cycle phase*** | ***Fertility*** | ***Ethnicity*** | ***Collection site*** |
| --- | --- | --- | --- | --- | --- | --- | --- | --- |
| **GSE232713** | Hiao-Yan Li | 33.93  (28-38) | Not provided | Not provided | Secretory (14) | Infertile (14) | Not provided | China |
| **GSE153739 &**  **GSE153740 ***** | Kashmira Bane | 31.70  (Not provided) | OE (4)  PE (3)  OE, PE (1) | II*  III*  IV* | Secretory (8)  Proliferative (7) | Fertile*  Infertile* | Not provided | India |
| **GSE141549** | Michael Gabriel | 33.43  (21-48) | OE (15)  PE (32)  DE (17)  RE (6)  SLE (7) | I (18)  II (19)  III (21)  IV (61)  Missing (2) | Proliferative (53)  Secretory (81)  Menstrual (14)  Missing (1) | Not provided | Not provided | Finland |
| **GSE134056** | Sadia Akter | Not provided  (18-49) | Not provided | Not provided | Not provided | Not provided | Not provided | USA |
| **GSE25628**  ****** | Stefania Crispi | 31.5  (22-42) | Not provided | II (2)  IV (6) | Proliferative (8) | Not provided | Not provided | Italy |
| **GSE37837** | Meraj A Khan | 33.17  (24-40) | OE (18) | III (8)  IV (10) | Secretory (5)  Proliferative (13) | Fertile (18) | Not provided | India |
| **GSE6364** | Burney RO | 36.23  (22-50) | OE, PE (7), PE (2), OE, PE, RE (5)  RE, PE (7) | III*  IV* | Secretory (26)  Proliferative (11) | Fertile*  Infertile* | Caucasian (26), Asian (4),  Black (3), Asian, Indian (1)  Unknown (3) | USA |
| **GSE7305** | Aniko Hever | Not provided | OE (10) | Not provided | Secretory (6)  Proliferative (2)  Unknown (2) | Not provided | Caucasian (10) | USA |

**Supplementary Table S2.** Clinical characteristics of patients per dataset. Abbreviations in Type of endometriosis columns denote: Ovarian endometriosis, OE; Peritoneal endometriosis, PE; Deep endometriosis; Rectovaginal endometriosis, RE; Sacrouterine ligament endometriosis, SLE. *No specific information regarding number of patients. ** No clinical data for healthy patients (n=6); clinical data available for patients with endometriosis (n=8); for each affected woman, two biopsies were obtained for the eutopic and ectopic endometrium respectively, while only one biopsy from the healthy eutopic endometrium was collected. For GSE153739 and GSE153740, data of 15 samples is deposited in GEO whilst clinical data in the corresponding article is provided for all 66 patients.

| **A** | | | | |
| --- | --- | --- | --- | --- |
| **Gene ontology term** | **Gene count** | **%** | **P-Value** | **FDR** |
| ***Biological Processes (GOTERM_BP_DIRECT)*** |  | | | |
| cell adhesion | 65 | 6,6 | 1,9E-10 | 6,4E-07 |
| inflammatory response | 54 | 5,5 | 2,8E-10 | 6,4E-07 |
| positive regulation of angiogenesis | 29 | 2,9 | 7,7E-09 | 1,2E-05 |
| positive regulation of gene expression | 57 | 5,8 | 1,4E-08 | 1,6E-05 |
| kidney development | 23 | 2,3 | 7,9E-08 | 7,2E-05 |
| ***Molecular Functions (GOTERM_MF_DIRECT)*** |  | | | |
| protein binding | 759 | 76,7 | 1,9E-16 | 2,2E-13 |
| identical protein binding | 148 | 15 | 2,6E-10 | 1,5E-07 |
| extracellular matrix structural constituent | 25 | 2,5 | 4,8E-09 | 1,9E-06 |
| protein homodimerization activity | 72 | 7,3 | 2,3E-07 | 6,6E-05 |
| heparin binding | 27 | 2,7 | 1,3E-06 | 3,1E-04 |
| ***GO cell component (GOTERM_CC_DIRECT)*** |  | | | |
| extracellular exosome | 210 | 21,2 | 2,1E-22 | 1,4E-19 |
| extracellular region | 190 | 19,2 | 5,3E-17 | 1,7E-14 |
| extracellular space | 172 | 17,4 | 8,8E-16 | 1,9E-13 |
| cell surface | 80 | 8,1 | 1,3E-14 | 2,1E-12 |
| plasma membrane | 352 | 35,6 | 1,6E-11 | 2,1E-09 |
| **B** | | | | |
| **Gene ontology term** | **Gene count** | **%** | **P-Value** | **FDR** |
| ***Pathways (KEGG pathway)*** |  | | | |
| Complement and coagulation cascades | 25 | 2,5 | 5,7E-10 | 1,9E-07 |
| Staphylococcus aureus infection | 23 | 2,3 | 1,5E-07 | 2,5E-05 |
| Cell adhesion molecules | 27 | 2,7 | 8,6E-06 | 9,5E-04 |
| Human T-cell leukemia virus 1 infection | 33 | 3,3 | 1,6E-05 | 1,1E-03 |
| Hematopoietic cell lineage | 20 | 2 | 1,7E-05 | 1,1E-03 |
| ***Pathways (REACTOME pathway)*** |  | | | |
| Extracellular matrix organization | 51 | 5,2 | 3,2E-10 | 4,9E-07 |
| Regulation of Complement cascade | 15 | 1,5 | 8,2E-07 | 6,4E-04 |
| Complement cascade | 16 | 1,6 | 2,4E-06 | 1,3E-03 |
| Molecules associated with elastic fibres | 12 | 1,2 | 1,3E-05 | 4,3E-03 |
| Elastic fibre formation | 13 | 1,3 | 1,4E-05 | 4,3E-03 |

**Supplementary Table S3**. Top 5 enriched GO terms per Biological Processes, Molecular functions and Cell Component category (A) and KEGG and REACTOME pathways (B).

| GO | Description | Count | % | Log10(P) | Log10(q) |
| --- | --- | --- | --- | --- | --- |
| GO:0035239 | tube morphogenesis | 96 | 9.71 | -33.30 | -28.95 |
| GO:0048732 | gland development | 66 | 6.67 | -25.31 | -21.61 |
| GO:0071345 | cellular response to cytokine stimulus | 88 | 8.90 | -25.21 | -21.61 |
| GO:0009725 | response to hormone | 91 | 9.20 | -25.17 | -21.61 |
| GO:0008285 | negative regulation of cell population proliferation | 90 | 9.10 | -24.32 | -20.82 |
| GO:0048729 | tissue morphogenesis | 75 | 7.58 | -24.05 | -20.60 |
| GO:0008283 | cell population proliferation | 83 | 8.39 | -23.21 | -19.81 |
| GO:0006954 | inflammatory response | 72 | 7.28 | -22.44 | -19.14 |
| GO:0040017 | positive regulation of locomotion | 75 | 7.58 | -21.84 | -18.57 |
| M5884 | NABA CORE MATRISOME | 49 | 4.95 | -21.60 | -18.40 |
| GO:0022407 | regulation of cell-cell adhesion | 66 | 6.67 | -21.55 | -18.38 |
| R-HSA-1474244 | Extracellular matrix organization | 51 | 5.16 | -21.51 | -18.37 |
| GO:0030855 | epithelial cell differentiation | 75 | 7.58 | -21.42 | -18.31 |
| GO:0032103 | positive regulation of response to external stimulus | 70 | 7.08 | -21.21 | -18.12 |
| GO:0061061 | muscle structure development | 66 | 6.67 | -20.42 | -17.42 |

**Supplementary Table S4**. Top 15 clusters enriched in endometrial lesions with their representative enriched terms. Count refers to the number of genes from the user-provided lists that are associated with the specified ontology term. % represents the proportion of all user-supplied genes that are included in that ontology term (only genes with at least one ontology annotation are considered in the calculation). Log10(P) is the p-value expressed in base-10 logarithmic form. Log10(q) is the p-value adjusted for multiple testing, also in base-10 logarithmic form.

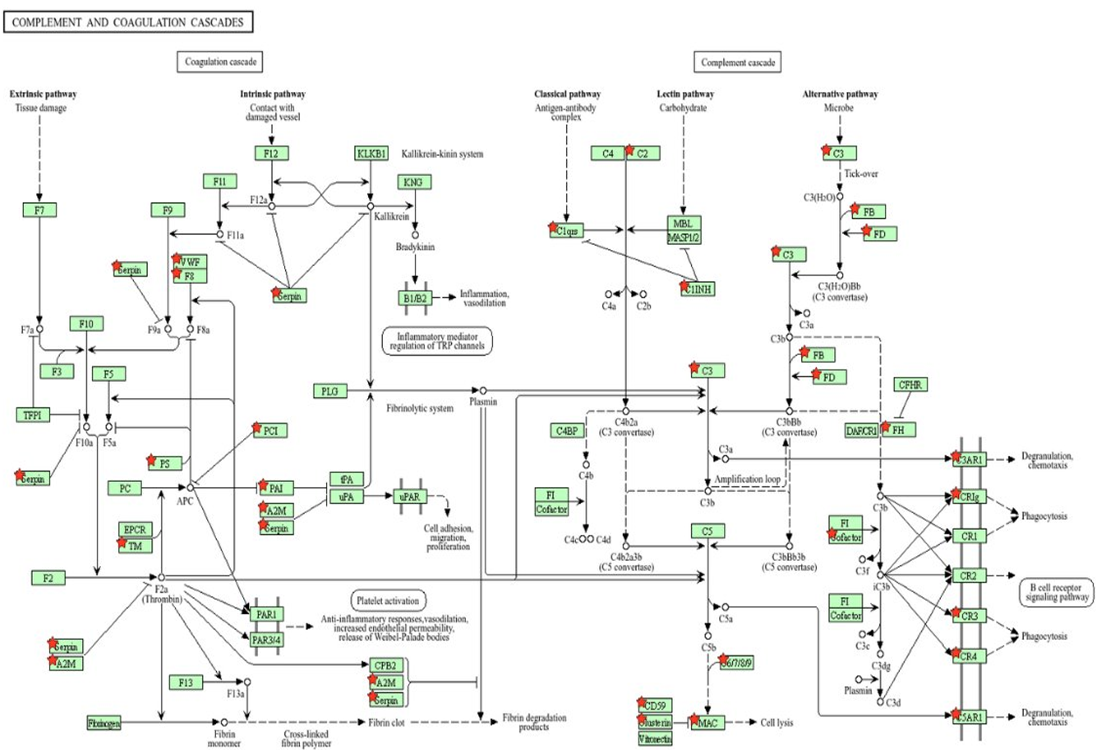

**Supplementary Figure S2**. Enrichment in the complement and coagulation pathway based on KEGG pathway analysis performed in DAVID. Pathway analysis against KEGG database showed that complement and coagulation cascades are the most enriched molecular events, red stars denote DEGs. Genes responsible for this enrichment include C2, C3, SERPINS and A2M.

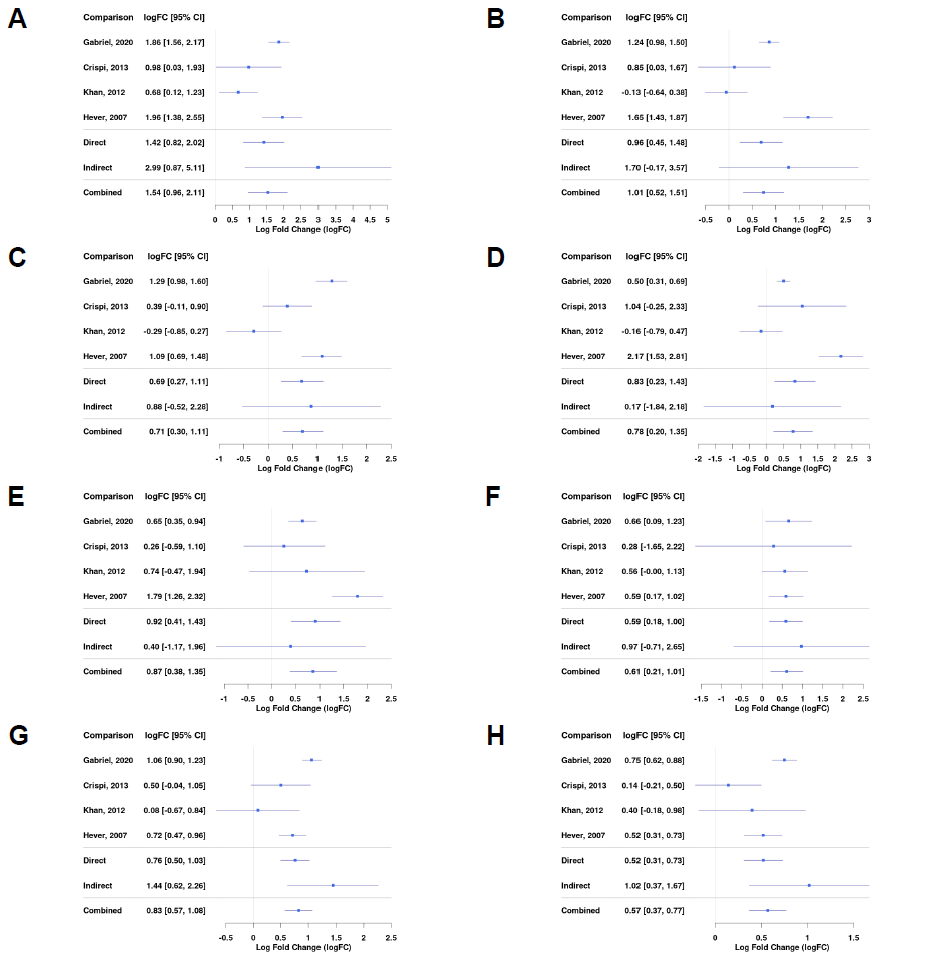

**Supplementary Figure S3**. **Differential gene expression across studies for selected genes including mast cells markers, JAK/STAT pathway and ECM.** Forest plot showing the expression of *CPA3 (mast cell carboxypeptidase A)* – A, *KIT (tyrosine-protein kinase KIT)* – B, *MS4A6A (Membrane Spanning 4-Domains A6A)* – C, *FCGR2B (Fc Gamma Receptor IIb)* – D, *S100A10 (S100 calcium-binding protein A10)* – E, *MMP-9 (Matrix metalloproteinase-9)* – F, *STAT5A (Signal Transducer And Activator Of Transcription 5A)* – G, *STAT5B (Signal Transducer And Activator Of Transcription 5B)* – H. Direct and indirect comparisons from meta-analysis are presented in row number five and six.
